# Circulating LIGHT (TNFSF14) and Interleukin-18 Levels in Sepsis-Induced Multi-Organ Injuries

**DOI:** 10.1101/2021.05.25.21257799

**Authors:** Hui-Qi Qu, James Snyder, John Connolly, Joseph Glessner, Charlly Kao, Patrick Sleiman, Hakon Hakonarson

## Abstract

The novel therapeutic target cytokine LIGHT (TNFSF14) was recently shown to play a major role in COVID19-induced acute respiratory distress syndrome (ARDS). This study aims to investigate the associations of plasma LIGHT and another potentially targetable cytokine, Interleukin-18 (IL-18), with ARDS, acute hypoxic respiratory failure (AHRF) or acute kidney injury (AKI), caused by non-COVID19 viral or bacterial sepsis. A cohort of 280 subjects diagnosed with sepsis, including 91 cases with sepsis triggered by viral infections, were investigated in this cohort study. Day 0 plasma LIGHT and IL-18, as well as 59 other biomarkers (cytokines, chemokines and acute-phase reactants) were measured by sensitive bead immunoassay and associated with symptom severity. We observed significantly increased LIGHT level in both bacterial sepsis patients (P=1.80E-05) and patients with sepsis from viral infections (P=1.78E-03). In bacterial sepsis, increased LIGHT level was associated with ARDS, AKI and higher Apache III scores, findings also supported by correlations of LIGHT with other biomarkers of organ failures. IL-18 levels were highly variable across individuals, and consistently correlated with Apache III scores, mortality, and AKI, in both bacterial and viral sepsis. There was no correlation between LIGHT and IL-18. For the first time, we demonstrate independent effects of LIGHT and IL-18 in septic organ failures. The association of plasma LIGHT with AHRF suggests that targeting the pathway warrants exploration, and ongoing trials may soon elucidate whether this is beneficial. Given the large variance of plasma IL-18 among septic subjects, targeting this pathway requires a precision application.

## 1. Introduction

The cytokine LIGHT (CD258), also known as tumor necrosis factor superfamily member 14 (TNFSF14), is a secreted protein of the TNF superfamily, recognized by the herpesvirus entry mediator (HVEM, also known as TNFRSF14), the lymphotoxin B receptor (LTβR, also known as TNFRSF3), and by decoy receptor, DcR3 (also known as TNFRSF6B) [1]. LIGHT exhibits inducible expression and competes with herpes virus glycoprotein D for binding to HVEM on T lymphocytes[2]. LIGHT is a ligand for TNFRSF14, which is a member of the tumor necrosis factor receptor superfamily, also known as HVEM ligand (HVEML). This protein functions as a costimulatory factor for the activation of lymphoid cells aimed at opposing infection by herpesvirus[2]. It additionally stimulates the proliferation of T cells, and triggers apoptosis of various tumor cells[3]. In addition to LIGHT, Interleukin-18 (IL-18) has been repeatedly highlighted as a major player in sepsis[4, 5]. As the interferon-γ inducing factor and a central inflammatory mediator, IL-18 has been demonstrated to play important roles in septic shock, organ failures, and sepsis mortality, suggesting a potential therapeutic target in sepsis[6-8]. Treatment with anti-IL-18 neutralizing antibodies has been shown to have significantly protective effects in a mouse model of sepsis [9].

Inflammatory reactions and immune responses are known to be dysregulated and compromised in patients with sepsis, the syndrome of infection complicated by organ injuries[10, 11]. In this regard, acute respiratory distress syndrome (ARDS) and acute kidney injury (AKI) are major complications of severe infections[12, 13], while the mechanisms of immune dysfunction causing these complications remains unknown. Sepsis typically manifests as an uncontrolled systemic inflammatory process, involving the microvasculature of multiple organ systems and manifesting as organ injury in one or multiple organs, including ARDS, AKI, disseminated intravascular coagulation (DIC)[14], or delirium[15, 16]. Liver, heart, and brain failures are also relatively common manifestations.

LIGHT, has recently come into the spotlight as a potent pro-inflammatory mediator and has been suggested as an important therapeutic target for immune regulation due to its central role in the function of activated T cells[17-19]. Previous study showed that soluble LIGHT induces proinflammatory changes in endothelial cells under systemic inflammatory activation[20]. The potential contribution of LIGHT to both inflammation and tissue fibrosis identifies it as a compelling potential contributor to ARDS and particularly fibroproliferative ARDS, which has very poor clinical outcomes. A recent study suggested an important role of LIGHT/HVEM expression in experimental lung injury in mice[21]. Nonetheless, it is worth attention that LIGHT may act as a double-edged sword. Due to its role in the innate immune response and inflammation recovery, *Tnfsf14*(-/-) mice develop more severe disease pathogenesis of intestinal inflammation[22]. For this reason, our working hypothesis is that LIGHT plays its detrimental effects in sepsis in binomial mode, i.e. only when its level is higher than a particular threshold. Among the LIGHT receptors, both HVEM and LTβR may drive the inflammatory response initiated by LIGHT[23]. In contrast, the decoy receptor DcR3 is able to neutralize the function of LIGHT[24]. Through this modulatory effect, our previous study suggested that loss of function of the DcR3 gene may be involved in the genetic risk of inflammatory bowel disease (IBD)[25]. High level of LIGHT beyond a threshold may also exceed the limit of DcR3 modulation.

Among other inflammatory biomarkers measured in this study, IL-18 is another pro-inflammatory LIGHT-like cytokine known to activate and regulate both innate and adaptive immunity, which we have an opportunity to modulate with an anti-IL18 neutralizing mAb. Its dysregulation has been shown to result in auto-inflammatory diseases[26, 27]. While IL-18 neutralizing mAb has been suggested for the treatment of various autoimmune and autoinflammatory diseases by experimental studies and clinical trails[28, 29], its role in sepsis is less clear.

We sought to investigate the potential role of LIGHT and IL18 in bacterial and viral-induced sepsis unrelated to COVID-19, and specifically the proteins’ associations with ARDS, acute hypoxic respiratory failure, and AKI as major sepsis complications in 280 patients with sepsis. Of those, 189 had either culture proven or presumed bacterial sepsis and 91 had PCR-diagnosed viral sepsis that resulted in hospitalization and admission to the intensive care unit (ICU). To contextualize the inflammatory cytokine milieu given the relative novelty of LIGHT, we also measured 59 inflammation or inflammasome biomarkers. This study highlights the significant roles of LIGHT and IL-18 in the severity of sepsis and for the first time, we demonstrate a key damaging role of LIGHT in patients with sepsis complicated by ARDS or multi-organ failures.

## 2. Materials and Methods

Subjects: This is a nested study with cases drawn from a molecular cohort study. Consecutive subjects (n=191) with sepsis enrolled to the Molecular Epidemiology of SepsiS in the ICU (MESSI) cohort study[30, 31] between 2016 and 2017 were included in this plasma investigation. As only 2 subjects during this time had viral sepsis and to enrich our population for viral sepsis given the reports of LIGHT associating with ARDS due to COVID-19, we broadened our time period from 2014 and 2019 to capture all subjects with viral sepsis during this time, which added 89 subjects. As the results, 189 (67.5%) cases diagnosed with bacterial sepsis and 91 (32.5%) cases diagnosed with PCR-confirmed viral sepsis were included in this study (Table 1). Each cohort was compared respectively with 22 controls from a random deidentified dataset of healthy (population based) subjects within comparable age, sex, and ethnic background to the sepsis cases. As has been previously published[30, 31], subjects were eligible for MESSI if, at the time of admission to the intensive care unit, they had strongly suspected infection and evidence of acute organ failure consistent with Sepsis-3; prior to 2016 we used the consensus definition for “severe sepsis” described in Sepsis-2[32]. We obtained cold residual plasma at day 0 from the blood collected clinically in the emergency room or on admission to the intensive care unit for subjects transferred from the floor. The clinical protocol is to spin citrated vacutainers upon receipt and to keep the plasma at 4°C after centrifuging; our research team collected the plasma at 12 – 36 hours after centrifuge, aliquoted, and froze the plasma at -80°C until assay.

**Table 1.** Clinical characteristics of the research subjects.

| **Clinical Information** | **Bacterial Sepsis (n=189)** | **Viral Sepsis (n=91)** |
| --- | --- | --- |
| **Age (median, 1<sup>st</sup> and 3<sup>rd</sup> quartiles)** | 61.5 (52.1, 71.4) | 61 (52.5, 69) |
| **sex** | 111(58.7%) males; 78(41.3%) females | 51(56.0%) males; 40(44.0%) females |
| **BMI (x ± s)** | 27.05±7.70 | 27.59±7.37 |
| **race** | White: 121; Black: 57; Asian: 8; Native American: 1; Other: 2 | White: 55; Black: 31; Asian: 2; Other: 3 |
| **ARDS** | Yes: 65(34.4%); No: 124(65.6%) | Yes: 36(39.6%); No: 55(60.4%) |
| **AHRF** | Yes: 71(37.8%); No: 117(62.2%) | Yes: 50(55.6%); No: 40(44.4%) |
| **AKI** | Yes: 114(61.6%); No: 71(38.4%) | Yes: 49(53.8%); No: 42(46.2%) |
| **APACHE III** | 101.6±36.9 | 90.8±39.4 |
| **LOS** | 19.38±20.78 | 17.70±19.98 |
| **Mortality** | Yes: 77(40.7%); No: 112(59.3%) | Yes: 33(36.3%); No: 58(63.7%) |

Phenotype determination: Subjects were enrolled prior to culture positivity and then all microbiologic data from 7 days prior to and 7 days after day 0 (ICU admission) were reviewed by physician investigators to classify primary and, if present, secondary body site of infection and to record the pathogens identified. The classification of ARDS required meeting oxygen and radiographic Berlin definition criteria on the same calendar day while receiving invasive ventilation[33]. We defined acute hypoxic respiratory failure (AHRF) as the patient manifesting hypoxia with arterial oxygen tension relative to fraction of inspired oxygen (PaO_2_:FiO_2_or PF ratio) ≤ 300 with or without mechanical ventilation. The determination of AKI was made in patients without preexisting end stage renal disease, based on serum creatinine using the Kidney Disease Improving Global Outcomes (KDIGO) guidelines[34]. To adjust for severity of illness at admission, we calculated the Acute Physiology and Chronic Illness III (APACHE III) score[35].

Measurement of circulating cytokines: We measured plasma LIGHT and IL-18, as well as an additional 59 biomarkers (cytokines, chemokines and acute-phase reactants), using the Quanterix high sensitivity single molecular array technology (Myriad RBM Simoa™ Services, Austin, TX), with LIGHT as a customized addition to the Human Inflammation MAP® v. 1.1. The list of the biomarkers measured is shown in Table S1.

Data analysis: The plasma levels of the measured biomarkers were normalized by natural log transformation for further analysis, due to the large dynamic ranges of the biomarkers. Elevated LIGHT level was defined as >2 standard deviations (SD) above mean in reference controls. Statistical analysis, including independent-samples t-test, One-Way ANOVA test, bivariate correlations, and non-parametric Chi-square test, were done by the IBM SPSS Statistics Version 23 software. Partial correlation was performed to control the effects of covariates. P values <0.001 are presented in scientific notation.

## 3. Results

### 3.1. Sepsis Population Characteristics

The study population included 280 subjects with sepsis, of whom 189 had sepsis of primarily bacterial origin and 91 had PCR-detected viral infection. Characteristics of the population are detailed in Table 1. Lung was the most common primary site of infection, accounting for 55% of sepsis, and 24% manifested bacteremia. For 5% of subjects, the infectious source remained unclear.

Among subjects with viral sepsis, 92% were pulmonary sepsis and influenza was the most common pathogen (62%) with additional viruses detected including respiratory syncytial virus, human metapneumovirus, adenovirus, non-SARS coronavirus, parainfluenza, rhinovirus/enterovirus, cytomegalovirus, and herpes simplex virus. Among the bacterial sepsis patients, 35% of culture proven cases were gram positive bacteria (most commonly Streptococcus, Staphylococcus, Enterococcus, and Gram-Positive Rod) and 55% were due to gram negative bacteria (most commonly Escherichia coli, Pseudomonas aeruginosa, Klebsiella pneumoniae, Acinetobacter baumannii, and gram-negative rod).

Pulmonary sepsis was strongly associated with AHRF (odds ratio, OR 3.29, 95% CI 1.98 – 5.47, p<0.001) and ARDS (OR 2.32, 95% CI 1.33 – 3.73, p=0.002). Viral sepsis was more likely to develop AHRF compared to non-viral sepsis (OR 1.96, 95% CI 1.18 – 3.24), p=0.009) but was not more likely to develop ARDS. Patients developed AKI, with similar rates between viral and non-viral sepsis, and 37% had died by day 30.

### 3.2. Increased plasma LIGHT levels in patients with sepsis

Increased LIGHT levels were observed in the sepsis cohort of 280 patients overall, compared to control levels from random adult samples, and this was attributed to increased LIGHT levels in both the bacterial and viral subsets of sepsis (independent t-test, Table 2). LIGHT levels were found to correlate inversely with age (r=-0.120, *P*=0.045), i.e. older age correlated with lower levels of LIGHT, whereas sex, race and BMI did not show correlation with LIGHT levels. Patients with viral sepsis showed trend of having lower LIGHT levels than patients with bacterial sepsis, though not statistically different (*P*=0.060). In addition, as shown in Table S3, several other biomarkers demonstrated association with organ failure in the overall sepsis group, including both bacterial and viral sepsis.

**Table 2.**
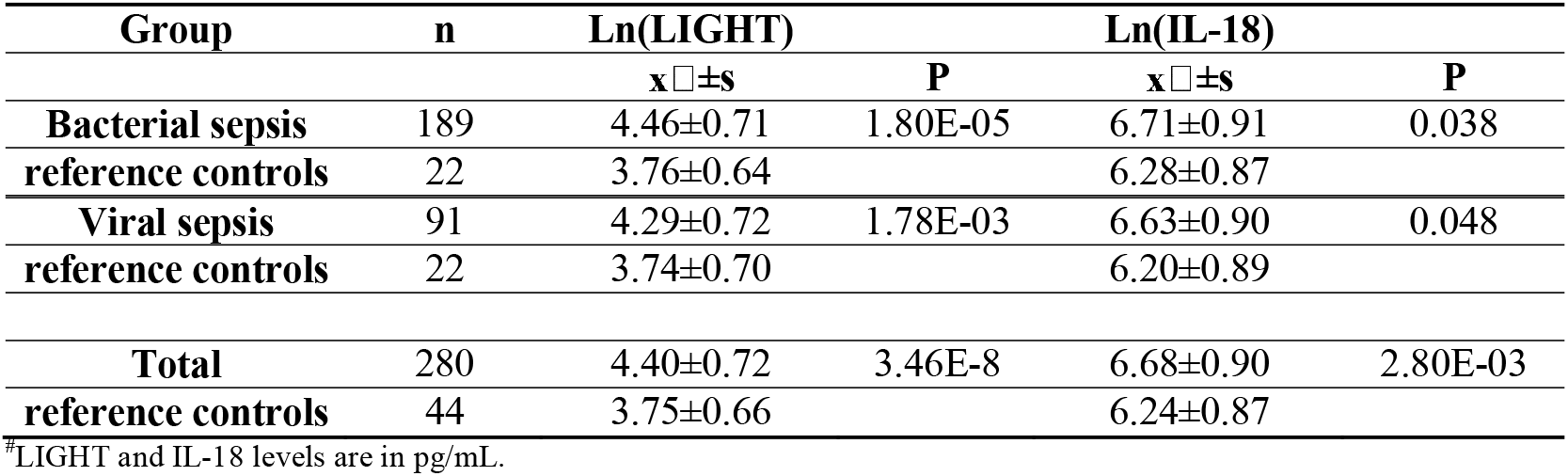
Ln(LIGHT) and Ln(IL-18) levels in bacterial and viral sepsis^#.^

| Group | n | Ln(LIGHT) |  | Ln(IL-18) |  |
| --- | --- | --- | --- | --- | --- |
| | | x ± s | P | x ± s | P |
| **Bacterial sepsis** | 189 | 4.46±0.71 | 1.80E-05 | 6.71±0.91 | 0.038 |
| **reference controls** | 22 | 3.76±0.64 | | 6.28±0.87 | |
| **Viral sepsis** | 91 | 4.29±0.72 | 1.78E-03 | 6.63±0.90 | 0.048 |
| **reference controls** | 22 | 3.74±0.70 | | 6.20±0.89 | |
| **Total** | 280 | 4.40±0.72 | 3.46E-8 | 6.68±0.90 | 2.80E-03 |
| **reference controls** | 44 | 3.75±0.66 | | 6.24±0.87 | |

<sup>#</sup>LIGHT and IL-18 levels are in pg/mL.

To examine the cutoff of abnormal LIGHT in an unbiased way, we run the association test scan with all the 189 sepsis cases against 22 controls. Due to potential batch effects between the bacterial sepsis cohort and the viral sepsis cohort, only the bacterial sepsis cohort was scanned. Shown by the results (Table S2), the cutoff of Ln(LIGHT) 4.875 has the least false positive rate (1/22) in controls. 138 cases (73%) have low LIGHT level by this definition. Additionally, we did an association scan for the best cutoff of LIGHT by its association with APACHE III scores. As shown by the results (Table S2), the cutoff 4.913 (142 cases, 75%) has the most statistical significance for the association between LIGHT and APACHE III (*P*=5.66e-05). As both of the cutoffs 4.875 and 4.913 are close to 5.043, i.e. 2 times of standard deviation of the reference controls for Ln(LIGHT), we thus defined abnormal LIGHT level as Ln(LIGHT)>5.043. 40 (21.2%) bacterial sepsis cases had elevated LIGHT by this definition.

In the 91 patients with viral sepsis, 12 (13.2%) cases had increased LIGHT. In the 189 bacterial sepsis cases, 40 (21.2%) cases had increased LIGHT, and 1/22 (4.5%) controls had elevated LIGHT. Overall, binary increased LIGHT was associated with AHRF with r=0.123, P=0.043, and trend towards elevated Apache III score with r=0.114, P=0.060, also associating with age. However, this analysis is underpowered.

In the viral sepsis cases, the Apache III scores were lower compared to bacterial sepsis, fewer patients had elevated LIGHT and correlation of LIGHT with Apache III score, ARDS, AHRF, AKI, and mortality, was not significant. Borderline significance was seen between increased length of ICU/hospital stay (LOS) and elevated LIGHT (r=0.196, P=0.063). Stratified by hospital mortality, the inverse correlation was from survival patients (r=0.297, P=0.023), i.e. extended length of hospitalization in survival patients with elevated LIGHT.

In contrast, in the bacterial sepsis cases, elevated LIGHT was correlated with Apache III, with r=0.172, p=0.020; associated with ARDS, with OR(95% CI)= 2.180 (1.077, 4.414), p=0.028; associated with AHRF, with OR(95% CI)= 2.037 (1.011, 4.106), p=0.044; and associated with AKI, with OR(95% CI)= 2.179 (0.991, 4.791), p=0.049. In addition, we also observed inverse correlation between LIGHT concentrations and LOS (β=-0.153, p=0.039). Stratified by hospital mortality, the inverse correlation was from patients with hospital mortality, i.e. the higher LIGHT concentration, the sooner death (β=-0.235, p=0.040).

Taken together, these results suggest that LIGHT may be a stronger pathogenic driver in bacterial sepsis than in viral sepsis, and given that both Apache III scores and mortality rates were higher in the bacterial sepsis lend support for elevated Apache III scores be considered an approach to identify patients that may have most benefit from LIGHT neutralizing mAb therapy.

### 3.3. Variable plasma IL-18 levels in patients with sepsis

We observed a wide dynamic range of IL-18 plasma concentrations, from 80 to >32,400 pg/mL in bacterial sepsis; and 161 to 19,100 pg/mL in viral sepsis. Compared to plasma LIGHT levels, the increase of plasma IL-18 levels showed only nominal significance in patients with sepsis (Table 2). In our study, sex, age, and BMI did not show correlation with IL-18 levels. However, IL-18 levels were found to correlate with self-identified race, with African American subjects manifesting lower IL-18 levels than other populations (6.44±0.98 *vs*. 6.79±0.85, p=0.003). In this case, instead of defining abnormal IL-18 levels, we tested association of quantitative IL-18 levels with clinical phenotypes, controlled for race (Table 3). Overall, IL-18 levels were correlated with Apache III scores, mortality, and AKI, with highly significant p values; and correlated with AHRF and ARDS with nominal significance. In addition, the correlation of IL-18 with Apache III scores, mortality, and AKI, were consistently observed in both bacterial sepsis and viral sepsis. In contrast, the correlation of IL-18 with AHRF was only observed in viral sepsis. These results suggest that, unlike LIGHT with prominent effects mainly observed in bacterial sepsis, IL-18 has significant impact in both bacterial and viral sepsis. In further, considering that IL-18 can be neutralized by IL-18BP (Figure 1), we also tested the IL-18 effects corrected for IL-18BP (Table 3).

**Table 3.** Correlation of quantitative Ln(IL-18) levels with organ failures^#.^

| IL-18 | All cases |  | Bacterial sepsis |  | Viral sepsis |  |
| --- | --- | --- | --- | --- | --- | --- |
|  | r | p value | r | p value | r | p value |
| **ARDS** | 0.121* | 0.045 | 0.103 | 0.165 | 0.167 | 0.117 |
| **AHRF** | 0.145* | 0.017 | 0.101 | 0.175 | 0.251* | 0.018 |
| **AKI** | 0.187** | 0.002 | 0.151* | 0.041 | 0.257* | 0.015 |
| **Apache III** | 0.301** | 0.000 | 0.278** | 1.40E-04 | 0.340** | 0.001 |
| **LOS** | 0.034 | 0.577 | 0.064 | 0.392 | -0.035 | 0.743 |
| **Mortality** | 0.288** | 0.000 | 0.254** | 0.001 | 0.350** | 0.001 |
| *Corrected for IL-18BP by partial correlation* |  |  |  |  |  |  |
|  | r | p value | r | p value | r | p value |
| **ARDS** | 0.144* | 0.018 | 0.147* | 0.047 | 0.16 | 0.135 |
| **AHRF** | 0.163** | 0.007 | 0.15* | 0.043 | 0.228* | 0.033 |
| **AKI** | 0.091 | 0.135 | 0.084 | 0.262 | 0.127 | 0.24 |
| **Apache III** | 0.239** | 7.084E-05 | 0.223** | 0.003 | 0.272* | 0.01 |
| **LOS** | 0.085 | 0.162 | 0.142 | 0.056 | -0.023 | 0.829 |
| **Mortality** | 0.269** | 7.337E-06 | 0.242** | 0.001 | 0.321** | 0.002 |

<sup>#</sup>Controlled for race.

**Figure 1.**
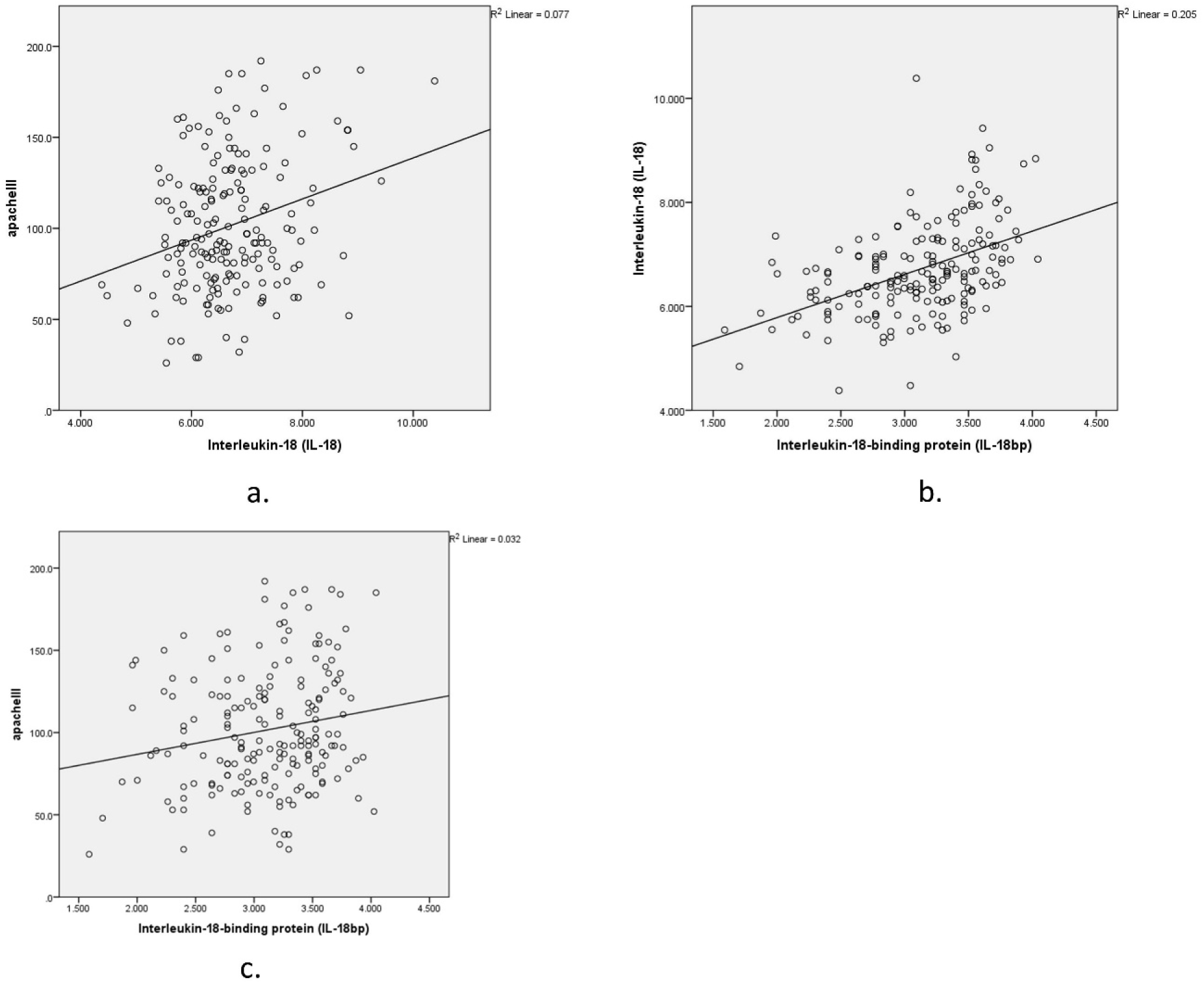
Correlations of Ln(IL-18), Ln(IL-18BP), and Apache III score in bacterial sepsis. The plots presented demonstrate the moderating effects of IL-18BP. To assess the correlation between IL-18 and Apache III score (a), the correlation between IL-18BP and IL-18 (b), as well as the potential correlation between IL-18BP and Apache III score (c), were controlled for in this study.

### 3.4. Other biomarkers in sepsis

The 59 exploratory biomarkers include a comprehensive list of inflammatory analytes and pathways integrated in the Human Inflammation MAP® v. 1.1 multiplexed immunoassays. Among the 59 exploratory biomarkers tested (i.e., markers other than LIGHT and IL-18, Table S3), the most significant associations included: Tissue Inhibitor of Metalloproteinases 1 (TIMP-1) was positively correlated with Apache III, with highly significant P values in both bacterial and viral sepsis; Plasminogen Activator Inhibitor 1 (PAI-1) was positively correlated with ARDS and AHRF in both bacterial and viral sepsis; and Tumor necrosis factor receptor 2 (TNFR2) was positively correlated with AKI in both bacterial and viral sepsis (Table 4). In bacterial sepsis, LIGHT was positively correlated with TIMP-1, PAI-1, and TNFR2. The above associations of LIGHT with these disease associated biomarkers are in keeping with the effects of LIGHT that we have observed in association with severe bacterial sepsis and its complications. IL-18 levels were correlated with disease biomarkers in both bacterial and viral sepsis. IL-18 levels were not correlated with elevated LIGHT levels in either bacterial or viral sepsis.

**Table 4.**
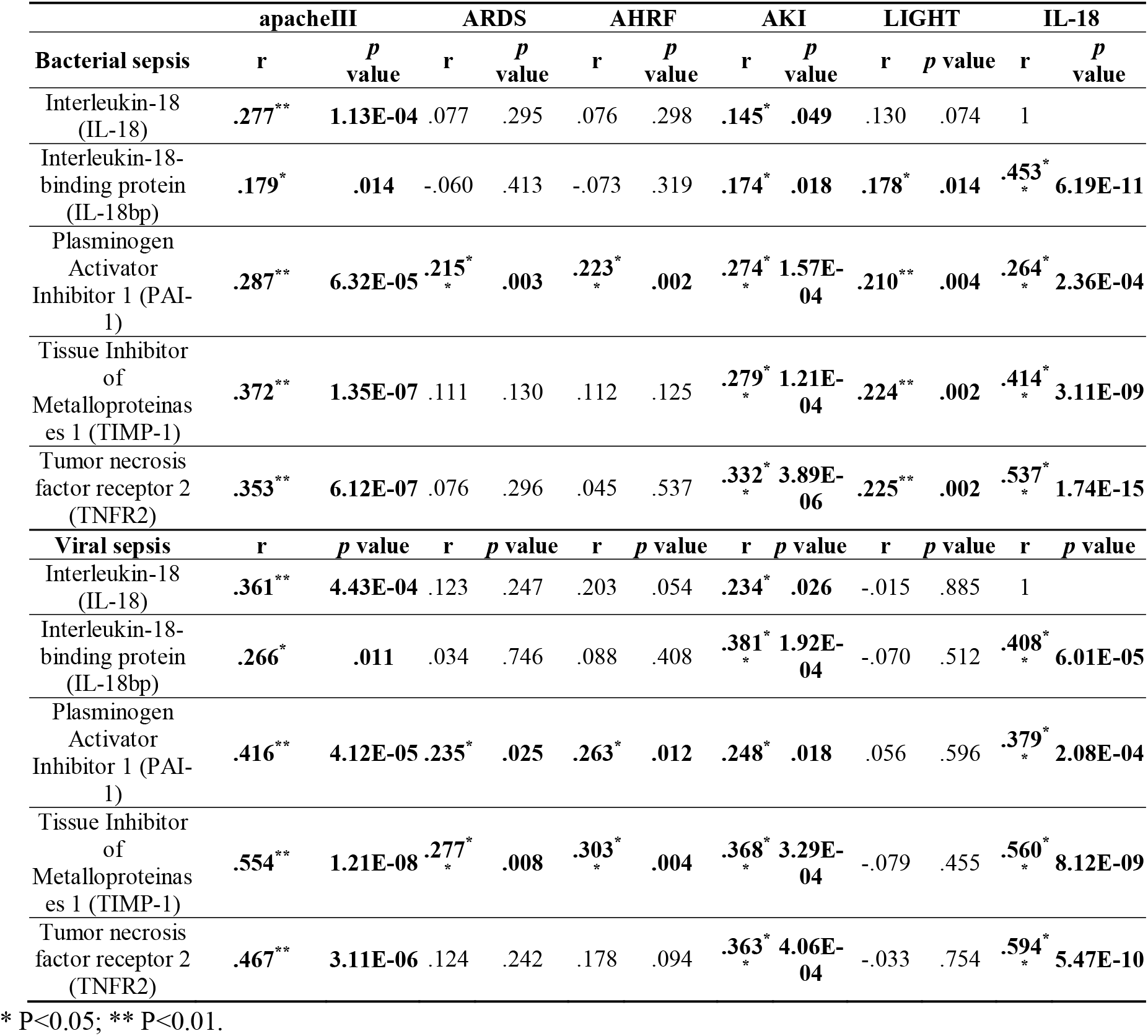
Correlation of elevated LIGHT and Ln(IL-18) with biomarkers of organ failures.

## 4. Discussion

### 4.1. LIGHT and IL-18 levels as biomarkers in sepsis

In this study, we report elevated LIGHT levels in both bacterial and viral sepsis. In bacterial sepsis, LIGHT is correlated with extended length of hospitalization, increased sepsis severity and sepsis complications. Viral sepsis cases had higher prevalence of ARDS and AHRF than bacterial sepsis. The lack of correlation of LIGHT in viral sepsis with Apache III score, ARDS, AHRF, and AKI is likely a reflection of smaller sample size (n=91).

In contrast to LIGHT as a mediator of immunoregulation, IL-18 is a proinflammatory cytokine, inducing the production of interferon-γ (IFNγ)[36]. Previous study has suggested IL-18 as a potential therapeutic target due to its crucial roles in sepsis[7]. In our study, IL-18 levels were consistently observed for correlations with increased sepsis severity and sepsis complications in both bacterial and viral sepsis. IL-18BP is the specific inhibitor of IL-18, and neutralizes IL-18 activities[36]. Previous study has shown that IL-18 mediates ischemic acute tubular necrosis in AKI[37], whereas our study shows that IL-18BP has much stronger correlation with AKI than IL-18 in sepsis. Apart for AKI, the IL-18 effects remain significant after correction for IL-18BP. In our study, IL-18 levels weren’t correlated with elevated LIGHT levels in either bacterial or viral sepsis, suggesting independent effects of LIGHT and IL-18 in sepsis rendering a combination therapy with neutralizing antibodies to LIGHT and IL18 a feasible choice.

### 4.2. LIGHT/IL-18 levels and Apache III score

The association of increased LIGHT level and Apache III score in bacterial sepsis, as well as highly significant correlation of IL-18 with Apache III score in both bacterial and viral sepsis, observed in this study may be explained in part by the association of LIGHT/IL-18 with increased risk of ARDS, AHRF, and AKI.

TIMP-1 is the most significant biomarker of Apache III score in both bacterial and viral sepsis as shown in this study. TIMP-1 is a key regulator of degradation of extracellular matrix. The matrix metalloproteinases (MMPs) degrade extracellular matrix and modulate cytokine and chemokine activity, which play important roles in immune responses to infection[38]. TIMP-1 is a natural inhibitor of MMPs[39]. TIMP-1 also functions as a cytokine, and promotes cell proliferation and has anti-apoptotic function[40, 41]. The study by Lorente et al. showed increased level of TIMP-1 as a biomarker to predict the clinical outcome of patients with sepsis[42].

Significantly increased expression of TIMP-1 by activated leukocytes has been observed in inflammatory tissues [43] [44]. Shown by our study, LIGHT is positively associated with TIMP-1 in bacterial sepsis, which may be related to its roles in the function of activated T cells[17-19] and systemic inflammatory activation[20]. IL-18 is consistently correlated with TIMP-1 with highly statistical significance in both bacterial and viral sepsis (Table 4). We observed positive correlation between IL-18 and IL-6/IL-10 in both both bacterial and viral sepsis (Table S3). Previous studies have shown that IL-6 induces the synthesis of TIMP-1[45], and IL-10 enhances release of TIMP-1 from peripheral blood mononuclear cells (PBMC)[46].

### 4.3. LIGHT/IL-18 levels and ARDS/AHRF

PAI-1 levels show significant correlation with ARDS and AHRF, which are also positively correlated with LIGHT levels in bacterial sepsis; and positively correlated with IL-18 levels in both bacterial and viral sepsis. PAI-1 is the main physiological plasminogen activator inhibitor, which is critical for regulating the fibrinolytic system and maintaining normal hemostasis[47]. Disseminated intravascular coagulation (DIC) is an important pathogenesis in early stage of acute lung injury (ALI) and ARDS[48]. Due to the crucial role of the fibrinolytic system and DIC in the pathophysiology of sepsis, PAI-1 levels have been shown to be important prognostic biomarker in sepsis[49]. Elevated in lung injury, alveolar PAI-1 levels can also predict ARDS in aspiration pneumonitis[50]. Encoded by the serpin family E member 1 gene (*SERPINE1*), the expression of PAI-1 is upregulated by LIGHT[51]. Therefore, the correction between LIGHT/ IL-18 levels and PAI-1 levels identified by this study suggests that LIGHT/ IL-18 may contribute to ARDS/AHRF in sepsis by increasing the PAI-1 levels.

### 4.4. LIGHT/IL-18 levels and AKI

In our study, TNFR2 is significantly correlated with AKI in both bacterial and viral sepsis. TNFR2 is the second receptor of the cytokines TNF and lymphotoxin-α, shown to mediate both proinflammatory and anti-inflammatory effects[52]. In recent years, TNFR2 has attracted research attentions as an emerging drug target for autoimmune diseases and cancer[53]. LIGHT levels in bacterial sepsis and IL-18 levels in both bacterial and viral sepsis were positively correlated with TNFR2. Both LIGHT and TNFR2 are integral components of the lymphotoxin system[17]. TNFR2 is encoded by the TNF receptor superfamily member 1B gene (*TNFRSF1B*) with restricted expression, e.g. in endothelial cells (ECs)[54] and lymphoid cells[55]. In the immune system, TNFR2 is predominantly expressed by a subset of highly suppressive CD4+CD25+FoxP3+ regulatory T cells (Treg) upon T-cell receptor (TCR) activation [56, 57]. The correlation of LIGHT with increased TNFR2 levels may be due to the suportive effects of LIGHT on the expansion and function of Tregs[58], while IL-18 has been shown of effect in driving Treg differentiation[59].

In sepsis, TNFR2 has been shown to associate with CD4+ T-cell impairment and post-septic immunosuppression by activation of Tregs[60]. Besides immunosuppression, significant up-regulation of TNFR2 in kidney injury was observed on tubular epithelial cells, including both distal convoluted tubules and proximal convoluted tubules[61]. Renal-expressed TNFR2 promotes renal monocyte recruitment by the IRF1 and IFN-β autocrine signaling[54], which may contribute to renal injury in sepsis. LIGHT expression by mucosal T cells has been shown of effects to promote TNFR2 expression in inflammatory tissue[62]. As the interferon-γ inducing factor, IL-18 may increase the expression of TNFR2 in kidney through interferon-γ, and has been shown of stimulation of renal epithelial cells to express TNFR2[63].

## 5. Conclusions

This study demonstrates independent associations of LIGHT and IL-18 in septic organ failures. Our study highlights two potential therapeutic targets in sepsis, i.e. the significantly increased LIGHT levels and the highly variable IL-18 levels across a subset of patients with sepsis. The detrimental roles of LIGHT in multi-organ failures were mainly seen in bacterial sepsis, including ARDS/AHRF, AKI, and its association with higher Apache III score as well as length of hospital stay with significant trend towards higher mortality rates. Consistently, the detrimental effects of LIGHT in bacterial sepsis were supported by the correlations of LIGHT with other biomarkers of organ failure suggesting a key role for LIGHT as an inflammatory driver of other detrimental mediators[64]. Given that LIGHT presents an interesting therapeutic target for severe inflammatory conditions, our results suggest for the first time that anti-LIGHT therapy with neutralizing mAb may be effective in a subset of patients with sepsis unrelated to COVID-19 infections. In contrast, IL-18 was associated with the detrimental outcome of both bacterial and viral sepsis, including increased mortality, and additionally supported by the correlations of IL-18 with other biomarkers involved with organ failure. The observed significant variance in IL-18 levels between individuals with sepsis highlights its potential as a precision therapeutic target with neutralizing mAb, by selecting patients with significantly elevated IL-18. The lack of correlation between LIGHT and IL-18 levels, as well as different correlations with other biomarkers, suggests independent and distinct roles of LIGHT and IL-18 in sepsis and that therapy directed against both of these cytokines should be given a consideration.

## Data Availability

Data available upon request.

## Supplementary Materials

Table S1: Sixty-one Inflammation biomarkers investigated in this study, Table S2: Scanning for LIGHT cutoff by association tests, Table S3: Correlation of elevated LIGHT and Ln(IL-18) with biomarkers of organ failures.

## Author Contributions

HH and HQQ designed and wrote the manuscript. HQQand PS contributed to data analysis and interpretation. JS, JC, JG, CK, and PS contributed by revising the manuscript and overall editing. HH contributed by supervision.

## Institutional Review Board Statement

This study was exempted by the Institutional Review Board (IRB) of the Children’s Hospital of Philadelphia. Human participants and person information are inaccessible to the research group.

## Informed Consent Statement

All human subjects or their proxies provided written informed consent.

## Data Availability Statement

Supporting data from this study can be obtained by emailing the corresponding author Dr. Hakon Hakonarson.

## Acknowledgments

Plasma samples and clinical phenotypes were graciously provided from the Molecular Epidemiology of SepsiS in the ICU (MESSI) cohort study at the Hospital of the University of Pennsylvania (PI: Nuala Meyer).

