## Supplementary material for "Circulating LIGHT (TNFSF14) and Interleukin-18 Levels in Sepsis-Induced Multi-Organ Injuries": Table S3

**Table S3** Correlation of elevated LIGHT and Ln(IL-18) with biomarkers of organ failures.#

|  | **apacheIII** | | **ARDS** | | **AHRF** | | **AKI** | | **LOS** | | **Mortality** | | **LIGHT** | | **IL-18** | |
| --- | --- | --- | --- | --- | --- | --- | --- | --- | --- | --- | --- | --- | --- | --- | --- | --- |
| **Bacterial sepsis** | **r** | ***p*  value** | **r** | ***p*  value** | **r** | ***p*  value** | **r** | ***p*  value** | **r** | ***p*  value** | **r** | ***p* value** | **r** | ***p* value** | **r** | ***p*  value** |
| T-Cell-Specific Protein RANTES (RANTES) | **-.160^*^** | **.028** | -.103 | .160 | -.088 | .230 | -.017 | .823 | -.054 | .457 | **-.226^**^** | **.002** | .112 | .125 | **-.331^**^** | **3.26E-06** |
| Apolipoprotein(a) (Lp(a)) | **-.213^**^** | **.003** | -.106 | .148 | -.101 | .170 | **-.223^**^** | **.002** | .060 | .409 | **-.312^**^** | **1.27E-05** | -.105 | .152 | **-.304^**^** | **2.06E-05** |
| Beta-2-Microglobulin (B2M) | **.282^**^** | **8.25E-05** | .094 | .200 | .053 | .474 | **.321^**^** | **8.27E-06** | -.094 | .201 | **.197^**^** | **.007** | **.157^*^** | **.031** | **.320^**^** | **7.04E-06** |
| Complement C3 (C3) | **-.271^**^** | **1.63E-04** | **-.155^*^** | **.034** | **-.157^*^** | **.032** | -.141 | .055 | .113 | .123 | **-.206^**^** | **.005** | -.097 | .185 | **-.198^**^** | **.006** |
| Eotaxin-1 | **.181^*^** | **.013** | -.032 | .666 | -.001 | .985 | .036 | .630 | **-.151^*^** | **.039** | **.146^*^** | **.044** | .080 | .272 | **.159^*^** | **.029** |
| Factor VII | **-.259^**^** | **3.23E-04** | -.087 | .235 | -.085 | .245 | -.135 | .066 | **.150^*^** | **.039** | **-.312^**^** | **1.22E-05** | **-.190^**^** | **.009** | **-.169^*^** | **.020** |
| Ferritin (FRTN) | **.214^**^** | **.003** | -.013 | .863 | -.011 | .883 | .076 | .305 | .130 | .075 | **.186^*^** | **.010** | .014 | .848 | **.541^**^** | **8.90E-16** |
| Fibrinogen | **-.167^*^** | **.021** | **-.204^**^** | **.005** | **-.188^**^** | **.010** | -.141 | .055 | .073 | .317 | **-.207^**^** | **.004** | -.050 | .495 | **-.199^**^** | **.006** |
| Interleukin-1 beta (IL-1 beta) | **.143^*^** | **.049** | .024 | .741 | .045 | .543 | .095 | .199 | -.056 | .442 | -.030 | .680 | -.008 | .908 | .027 | .711 |
| Interleukin-1 receptor antagonist (IL-1ra) | **.203^**^** | **.005** | .077 | .295 | .088 | .228 | .132 | .074 | **-.161^*^** | **.027** | .048 | .511 | **.155^*^** | **.033** | **.258^**^** | **3.43E-04** |
| Interleukin-6 (IL-6) | **.251^**^** | **4.92E-04** | .000 | .997 | -.026 | .725 | .123 | .095 | -.057 | .438 | .126 | .084 | .090 | .218 | **.156^*^** | **.032** |
| Interleukin-8 (IL-8) | **.239^**^** | **.001** | .014 | .853 | .006 | .937 | .103 | .162 | -.056 | .446 | **.166^*^** | **.022** | .093 | .205 | **.241^**^** | **.001** |
| Interleukin-10 (IL-10) | **.183^*^** | **.012** | .033 | .656 | .034 | .645 | .078 | .293 | -.070 | .335 | .066 | .368 | .086 | .239 | **.257^**^** | **3.54E-04** |
| Interleukin-18 (IL-18) | **.277^**^** | **1.13E-04** | .077 | .295 | .076 | .298 | **.145^*^** | **.049** | .084 | .250 | **.271^**^** | **1.61E-04** | .130 | .074 | 1 |  |
| Interleukin-18-binding protein (IL-18bp) | **.179^*^** | **.014** | -.060 | .413 | -.073 | .319 | **.174^*^** | **.018** | -.116 | .111 | .097 | .186 | **.178^*^** | **.014** | **.453^**^** | **6.19E-11** |
| Macrophage Inflammatory Protein-1 beta (MIP-1 beta) | **.172^*^** | **.018** | -.016 | .828 | -.024 | .742 | .079 | .284 | -.141 | .053 | .135 | .063 | .130 | .075 | **.227^**^** | **.002** |
| Matrix Metalloproteinase-3 (MMP-3) | **.195^**^** | **.007** | -.021 | .772 | -.051 | .485 | **.309^**^** | **1.83E-05** | -.023 | .751 | **.164^*^** | **.024** | -.017 | .818 | **.154^*^** | **.035** |
| Myoglobin | **.266^**^** | **2.24E-04** | .106 | .149 | .068 | .353 | **.270^**^** | **2.09E-04** | **-.180^*^** | **.014** | .122 | .096 | .122 | .095 | .105 | .151 |
| Plasminogen Activator Inhibitor 1 (PAI-1) | **.287^**^** | **6.32E-05** | **.215^**^** | **.003** | **.223^**^** | **.002** | **.274^**^** | **1.57E-04** | -.098 | .178 | **.264^**^** | **2.42E-04** | **.210^**^** | **.004** | **.264^**^** | **2.36E-04** |
| Serum Amyloid P-Component (SAP) | **-.169^*^** | **.020** | -.124 | .089 | -.128 | .079 | -.118 | .110 | .109 | .136 | **-.264^**^** | **2.46E-04** | -.080 | .272 | -.090 | .219 |
| Stem Cell Factor (SCF) | .121 | .098 | -.118 | .105 | **-.148^*^** | **.042** | **.330^**^** | **4.62E-06** | .036 | .618 | .070 | .342 | -.054 | .457 | **.152^*^** | **.037** |
| Thyroxine-Binding Globulin (TBG) | **-.218^**^** | **.003** | -.111 | .130 | -.127 | .082 | **-.152^*^** | **.039** | -.067 | .361 | **-.215^**^** | **.003** | .023 | .758 | -.045 | .541 |
| Tissue Inhibitor of Metalloproteinases 1 (TIMP-1) | **.372^**^** | **1.35E-07** | .111 | .130 | .112 | .125 | **.279^**^** | **1.21E-04** | -.092 | .207 | **.298^**^** | **3.05E-05** | **.224^**^** | **.002** | **.414^**^** | **3.11E-09** |
| Tumor necrosis factor receptor 2 (TNFR2) | **.353^**^** | **6.12E-07** | .076 | .296 | .045 | .537 | **.332^**^** | **3.89E-06** | **-.156^*^** | **.032** | **.298^**^** | **3.14E-05** | **.225^**^** | **.002** | **.537^**^** | **1.74E-15** |
| Vascular Cell Adhesion Molecule-1 (VCAM-1) | **.330^**^** | **3.52E-06** | .048 | .512 | .033 | .648 | **.238^**^** | **.001** | -.051 | .488 | **.310^**^** | **1.38E-05** | .114 | .117 | **.452^**^** | **6.42E-11** |
| Vitamin D-Binding Protein (VDBP) | **-.259^**^** | **3.24E-04** | -.065 | .372 | -.055 | .453 | **-.158^*^** | **.032** | **.156^*^** | **.032** | **-.302^**^** | **2.40E-05** | -.030 | .681 | -.099 | .176 |
| von Willebrand Factor (vWF) | **.247^**^** | **.001** | .055 | .456 | .024 | .746 | **.206^**^** | **.005** | -.037 | .608 | **.235^**^** | **.001** | **.156^*^** | **.032** | **.281^**^** | **9.03E-05** |
| **Viral sepsis** | **r** | ***p* value** | **r** | ***p* value** | **r** | ***p* value** | **r** | ***p* value** | **r** | ***p* value** | **r** | ***p* value** | **r** | ***p* value** | **r** | ***p* value** |
| T-Cell-Specific Protein RANTES (RANTES) | **-.486^**^** | **1.03E-06** | **-.230^*^** | **.028** | -.137 | .198 | **-.250^*^** | **.017** | -.080 | .450 | **-.435^**^** | **1.63E-05** | **.255^*^** | **.015** | **-.439^**^** | **1.31E-05** |
| Apolipoprotein(a) (Lp(a)) | -.186 | .077 | -.135 | .203 | -.038 | .720 | -.104 | .327 | .010 | .924 | -.164 | .120 | -.146 | .167 | **-.226^*^** | **.031** |
| Beta-2-Microglobulin (B2M) | .193 | .066 | -.011 | .915 | .057 | .595 | **.441^**^** | **1.20E-05** | -.063 | .552 | .056 | .601 | -.118 | .267 | **.294^**^** | **.005** |
| Complement C3 (C3) | **-.466^**^** | **3.27E-06** | -.196 | .062 | -.099 | .352 | -.166 | .115 | .025 | .814 | **-.237^*^** | **.024** | .050 | .638 | **-.223^*^** | **.034** |
| Eotaxin-1 | **.292^**^** | **.005** | .059 | .575 | -.039 | .712 | .054 | .612 | .035 | .741 | .070 | .507 | .098 | .355 | .148 | .162 |
| Factor VII | **-.404^**^** | **7.18E-05** | **-.243^*^** | **.021** | -.177 | .094 | -.030 | .774 | .131 | .216 | **-.375^**^** | **2.53E-04** | -.062 | .558 | -.158 | .135 |
| Ferritin (FRTN) | **.383^**^** | **1.82E-04** | .109 | .306 | .123 | .247 | **.241^*^** | **.022** | .005 | .959 | **.401^**^** | **8.04E-05** | -.122 | .249 | **.551^**^** | **1.57E-08** |
| Fibrinogen | **-.252^*^** | **.016** | **-.230^*^** | **.028** | -.085 | .426 | .072 | .495 | .143 | .175 | -.161 | .127 | -.124 | .243 | -.105 | .321 |
| Interleukin-1 beta (IL-1 beta) | **.226^*^** | **.031** | **.263^*^** | **.012** | .187 | .077 | .092 | .386 | -.079 | .457 | **.236^*^** | **.024** | -.102 | .336 | .179 | .089 |
| Interleukin-1 receptor antagonist (IL-1ra) | **.461^**^** | **4.25E-06** | **.301^**^** | **.004** | **.267^*^** | **.011** | .173 | .102 | -.037 | .731 | .175 | .096 | -.011 | .916 | **.312^**^** | **.003** |
| Interleukin-6 (IL-6) | **.503^**^** | **3.68E-07** | **.347^**^** | **.001** | **.359^**^** | **.001** | .175 | .097 | .009 | .931 | **.296^**^** | **.004** | -.023 | .832 | **.304^**^** | **.003** |
| Interleukin-8 (IL-8) | **.562^**^** | **6.80E-09** | **.297^**^** | **.004** | **.244^*^** | **.021** | **.257^*^** | **.014** | .103 | .332 | **.428^**^** | **2.26E-05** | -.134 | .204 | **.529^**^** | **6.80E-08** |
| Interleukin-10 (IL-10) | **.593^**^** | **5.94E-10** | **.377^**^** | **2.27E-04** | **.263^*^** | **.012** | **.226^*^** | **.032** | -.099 | .348 | **.328^**^** | **.002** | -.051 | .631 | **.468^**^** | **2.88E-06** |
| Interleukin-18 (IL-18) | **.361^**^** | **4.43E-04** | .123 | .247 | .203 | .054 | **.234^*^** | **.026** | .045 | .674 | **.408^**^** | **5.93E-05** | -.015 | .885 | 1 |  |
| Interleukin-18-binding protein (IL-18bp) | **.266^*^** | **.011** | .034 | .746 | .088 | .408 | **.381^**^** | **1.92E-04** | .000 | .998 | .183 | .082 | -.070 | .512 | **.408^**^** | **6.01E-05** |
| Macrophage Inflammatory Protein-1 beta (MIP-1 beta) | **.352^**^** | **.001** | .198 | .060 | .146 | .170 | .197 | .062 | .012 | .910 | **.292^**^** | **.005** | -.002 | .987 | **.503^**^** | **3.65E-07** |
| Matrix Metalloproteinase-3 (MMP-3) | **.244^*^** | **.020** | -.054 | .611 | .004 | .969 | **.228^*^** | **.030** | -.030 | .776 | .041 | .696 | -.112 | .290 | .141 | .182 |
| Myoglobin | **.346^**^** | **.001** | .081 | .444 | .033 | .759 | **.319^**^** | **.002** | **-.250^*^** | **.017** | .065 | .542 | -.203 | .054 | .154 | .146 |
| Plasminogen Activator Inhibitor 1 (PAI-1) | **.416^**^** | **4.12E-05** | **.235^*^** | **.025** | **.263^*^** | **.012** | **.248^*^** | **.018** | .054 | .614 | **.327^**^** | **.002** | .056 | .596 | **.379^**^** | **2.08E-04** |
| Serum Amyloid P-Component (SAP) | **-.527^**^** | **7.84E-08** | **-.342^**^** | **.001** | -.160 | .133 | -.167 | .114 | .004 | .968 | **-.323^**^** | **.002** | -.012 | .912 | **-.244^*^** | **.020** |
| Stem Cell Factor (SCF) | **.256^*^** | **.014** | -.092 | .384 | .001 | .994 | **.437^**^** | **1.46E-05** | .148 | .161 | .024 | .824 | -.140 | .185 | **.208^*^** | **.048** |
| Thyroxine-Binding Globulin (TBG) | **-.396^**^** | **1.04E-04** | -.077 | .471 | -.032 | .762 | -.178 | .091 | -.008 | .938 | **-.314^**^** | **.002** | .123 | .244 | -.144 | .175 |
| Tissue Inhibitor of Metalloproteinases 1 (TIMP-1) | **.554^**^** | **1.21E-08** | **.277^**^** | **.008** | **.303^**^** | **.004** | **.368^**^** | **3.29E-04** | -.008 | .943 | **.353^**^** | **.001** | -.079 | .455 | **.560^**^** | **8.12E-09** |
| Tumor necrosis factor receptor 2 (TNFR2) | **.467^**^** | **3.11E-06** | .124 | .242 | .178 | .094 | **.363^**^** | **4.06E-04** | .062 | .562 | **.307^**^** | **.003** | -.033 | .754 | **.594^**^** | **5.47E-10** |
| Vascular Cell Adhesion Molecule-1 (VCAM-1) | **.311^**^** | **.003** | .120 | .256 | .183 | .084 | **.340^**^** | **.001** | -.019 | .856 | **.261^*^** | **.012** | -.031 | .770 | **.550^**^** | **1.67E-08** |
| Vitamin D-Binding Protein (VDBP) | **-.411^**^** | **5.10E-05** | **-.318^**^** | **.002** | -.200 | .059 | -.108 | .309 | .077 | .471 | -.147 | .164 | -.076 | .476 | **-.216^*^** | **.040** |
| von Willebrand Factor (vWF) | **.234^*^** | **.025** | .058 | .583 | .055 | .604 | .144 | .174 | -.062 | .562 | .113 | .288 | -.104 | .325 | **.314^**^** | **.002** |

* P<0.05; ** P<0.01.

\# ***Summary of the biomarkers in sepsis***

(1) Biomarkers associated with Apache III score in both bacterial and viral sepsis cases

Among the 59 exploratory biomarkers tested (i.e., markers other than LIGHT and IL-18), TIMP-1, TNFR2, VCAM-1, PAI-1, IL-18BP, myoglobin, IL-6, vWF, IL-8, FRTN, IL-1Ra, MMP-3, IL-10, Eotaxin-1, MIP-1β, and IL-1β, were positively correlated with Apache III, in both bacterial and viral sepsis.

Interestingly, complement C3, Factor VII, VDBP, TBG, SAP, Fibrinogen, and RANTES, were all negatively correlated with Apache III score in both bacterial and viral sepsis.

(2) Biomarkers associated with ARDS and AHRF in both bacterial and viral sepsis

In addition to the correlation of LIGHT and IL-18 as shown above, among the 59 exploratory biomarkers, PAI-1 was positively correlated with ARDS and AHRF, whereas fibrinogen was negatively correlated with ARDS. Interestingly, another exploratory observation in patients with sepsis is that, IL-10, IL-6, IL-1Ra, IL-8, and TIMP-1, are associated with ARDS and AHRF in viral sepsis only at highly statistical significance. No such association was identified with bacterial sepsis in this study.

(3) Biomarkers associated with AKI in both bacterial and viral sepsis

In addition to the association of LIGHT and IL, among the 60 additional biomarkers, B2M, SCF, TNFR2, VCAM-1, TIMP-1, Myoglobin, MMP-3, PAI-1, IL-18 and IL-18BP, were positively correlated with AKI, indicative of worse clinical outcome.

(4) Biomarkers associated with elevated LIGHT levels

In viral sepsis, elevated LIGHT was positively correlated with RANTES. In bacterial sepsis, LIGHT was positively correlated with TIMP-1, TNFR2, PAI-1, IL-1Ra, vWF, IL-18BP, and B2M, while it was negatively correlated with Factor VII. The above associations of LIGHT with these disease associated biomarkers are in keeping with the effects of LIGHT that we have observed in association with severe bacterial sepsis and its complications.

(5) Biomarkers associated with IL-18 levels

IL-18 levels correlated with disease biomarkers in both bacterial and viral sepsis. In both bacterial and viral sepsis, IL-18 levels were positively correlated with Ferritin, IL-10, IL-18BP, PAI-1, TIMP-1, TNFR2, VCAM-1, IL-8, MIP-1β, vWF, IL-1Ra, IL-6, B2M, and SCF; and negatively correlated with RANTES, Lp(a), and C3. In bacterial sepsis, IL-18 levels were also positively correlated with Eotaxin-1 and MMP-3; and negatively correlated with Factor VII and Fibrinogen. In viral sepsis, IL-18 levels were also negatively correlated with SAP and VDBP. IL-18 levels were not correlated with elevated LIGHT levels in either bacterial or viral sepsis.
